# Hypothyroidism and its influence in the development of Metabolic Syndrome (MetS)

**DOI:** 10.1101/2021.03.13.21253240

**Authors:** D. J. L. L. Pinheiro, T. C. Santos, G. R. Spacini, A. P. S. Guerra, M. C. L Bacci, M. G Nemecek, J. R. C. Oliveira, R. L. M. Dantas, P. J. O. Cortez, J. Faber

**Affiliations:** Federal University of São Paulo–UNIFESP, Escola Paulista de Medicina, Department of Neurology and Neurosurgery, São Paulo, SP, Brasil; Federal University of São Paulo - UNIFESP, Institute of Science and Technology, Biomedical Engineering, São José dos Campos, SP, Brasil; Itajubá Faculty of Medicine, Magsul Clinic, Itajubá, MG, Brasil

**Keywords:** metabolic syndrome, hypothyroidism, comorbidities, exploratory analysis

## Abstract

**Background:** Metabolic syndrome (MetS) is a complex disorder that affects the cardiovascular system and it is generally associated with mental and metabolic health issues. To diagnose MetS it is essential to identify certain comorbidities related to cardiovascular diseases. Hypothyroidism, for instance, is a common disorder that reduces the basal metabolism, leading to the development of other diseases. Therefore, it is important to know how hypothyroidism can increase the chances of MetS.

**Objectives:** To assess whether there is an increased risk of patients with hypothyroidism developing MetS given the association of different comorbidities in a sample of an urban Brazilian population.

**Methods:** This was an observational, exploratory and retrospective study of patients diagnosed with hypothyroidism (using levothyroxine for hormonal reposition therapy).The incidence and prevalence of four specific comorbidities were quantified by histograms, and all comparisons were performed by using joint and conditional probabilities with their respective confident intervals.

**Results:** Consistent with the literature, our results also showed that there is a prevalence in women (79%) and in elderly patients (80%). Moreover, it also indicated that around 64% of the patients were women aged over 60 years and at least one comorbidity was associated with them. By analyzing the risk group for MetS, with two comorbidities, the combination with higher chances of acquiring a new comorbidity were Systemic arterial hypertension (SAH) and Diabetes Mellitus (DM), in this sample.

**Conclusion:** Our results indicate that hypothyroidism can increase the chances to develop MetS or may worsen its effects due to the association of common comorbidities present in both cases. Keywords: metabolic syndrome, hypothyroidism, comorbidities, statistical analysis.

## 1. Introduction

Hypothyroidism is a disease characterized by a deficit on thyroid hormone production, which can be severe or moderate (1). The diagnosis is defined biochemically by thyroid-stimulating hormone (TSH) evaluating if the concentration is lower the limit of the reference range. Therefore, the absence or excess of TSH, directly affects the metabolism associated with the systems they operate (2). Since hypothyroidism has no specific clinical symptoms and a pathognomonic sign to distinguish patients, the TSH is determinant (1). The normal range of TSH values, for individuals thyroid disease-free has been accepted as 0.45-4.5 mU/L (3). The levels accept to diagnosis the hypothyroidism change with the age and the American Association of Clinical Endocrinologists and National Academy of Clinical Biochemistry recommends measuring the levels of TSH for 2-3 month. If the level is not normalized, the patient is normally diagnosed with hypothyroidism (4). While, patients characterized with TSH deficit, generally undergo hormonal repositioning using levothyroxine (5,6,7).

Different studies have indicated that hypothyroidism is related or intensifies other clinical condition. Additionally, when patients are diagnosed with at least three components related to cardiovascular disease (CVD), such as hypertension, dyslipidemia, obesity and diabetes mellitus (8,9), they can be diagnosed with the Metabolic Syndrome (MetS) (10).

MetS is a major clinical challenge worldwide, usually caused due to sedentary lifestyle and unhealthy diet. It is a complex disorder that leads to several cardiovascular risks, mental abnormalities, and metabolic imbalances (10,11). Other studies have been reporting Hypothyroidism and MetS well-associated with risk factors for atherogenic cardiovascular disease (12,13). In these cases, it is pointed that patients with insulin resistance have common pathogenic mechanisms that cause the overlap between hypothyroid and MetS (14).

Most of the current investigations focus on the consequences of the MetS on the hypothyroidism (15,16,17). Here, we are interested to investigate the changes of a patient with hypothyroidism to develop MetS, given the combination of different comorbidities (inside a population of 327 patients). To evaluate it, we calculate all possible combinations related to the main prevalent comorbidities in this population: Systemic arterial hypertension (SAH), Obesity, Diabetes mellitus (DM), Dyslipidemia (DLP).

## 2. Methods

This project was approved by the Research Ethics Committee and it’s followed the ethical percepts of Brazilian Health National Council 466/2012, approval number: CAAE: 45972615.6.0000.5559, sight number 1.112.225. Medical records of 327 patients were collected for analysis. In our study, all patients were diagnosed with hypothyroidism and they were undergoing treatment with the medication levothyroxine sodium, with fantasy name’s Euthyrox, Levoid, Puram, Synthroid and Tiroidin.

### Data and Statistical analysis

Patient data was collected including age, gender, and body mass index (BMI), associated with the following categorical variables:

- Systemic arterial hypertension (SAH);
- Obesity (BMI > 30);
- Smoking;
- Diabetes mellitus (DM);
- Dyslipidemia (DLP);

The statistical analysis was carried out using MATLAB R2017b. The joint and conditional probability, associated with the prevalence and trends of comorbidities on the sample, for the diagnosis of metabolic syndrome, were performed by means of the patient’s clinical recordings.

Since MetS requires that the patient presents simultaneously at least 3 comorbidities, we calculate the chance that a patient with any 2 comorbidities have any third one. In this case, the conditional probability was calculated according to the rule: P(X|Y,Z) = P(X,Y,Z)/P(Y,Z), where X, Y and Z represent random variables associated with each one of the 4 comorbidities: Systemic arterial hypertension (SAH), Obesity, Diabetes mellitus (DM), Dyslipidemia (DLP), P(X,Y,Z) is the joint probability of a patient having 3 comorbidities simultaneously and P(X,Y) is the joint probability of a patient having 2 comorbidities simultaneously (18). The tabagism has been removed from this analysis because it does not fit into one of the risk diseases for diagnosing MetS.

All statistical significances were evaluated from the intersection of the probability confident intervals, according to 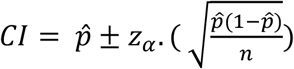, with *z*_*α*_=196, with *α* =5% (16). The skewness of the histograms was calculated following 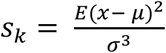.

### Limitations

this study is only observational with few samples that may compromise the stability of the claimed statistical relationships, requiring a more rigorous control study. However, we believe it is a preliminary evidence pointing to a possible relation between Hypothyroidism and Metabolic Syndrome.

## 3. Results

Figure 1 shows the age and BMI histograms, with (68.79 ± 10.05) for age, and (27.75 ± 4.84) for BMI. Particularly, Figure 1(a) shows a prevalence of older population (blue histogram), skewed to the right and spread out to the left, sk= -0.2602; and a concentration for no-obese patients (red histogram), sk= 0.9552, on this population. To complement the interpretation, figure 1 (b) shows that the relationship between the samples of men and women are undistinguished for age and BMI. The obesity is well emphasized by plotting age against BMI (since, obesity occurs when BMI>30), showing a high homogeneity related to these two classes.

**Figure 1.**
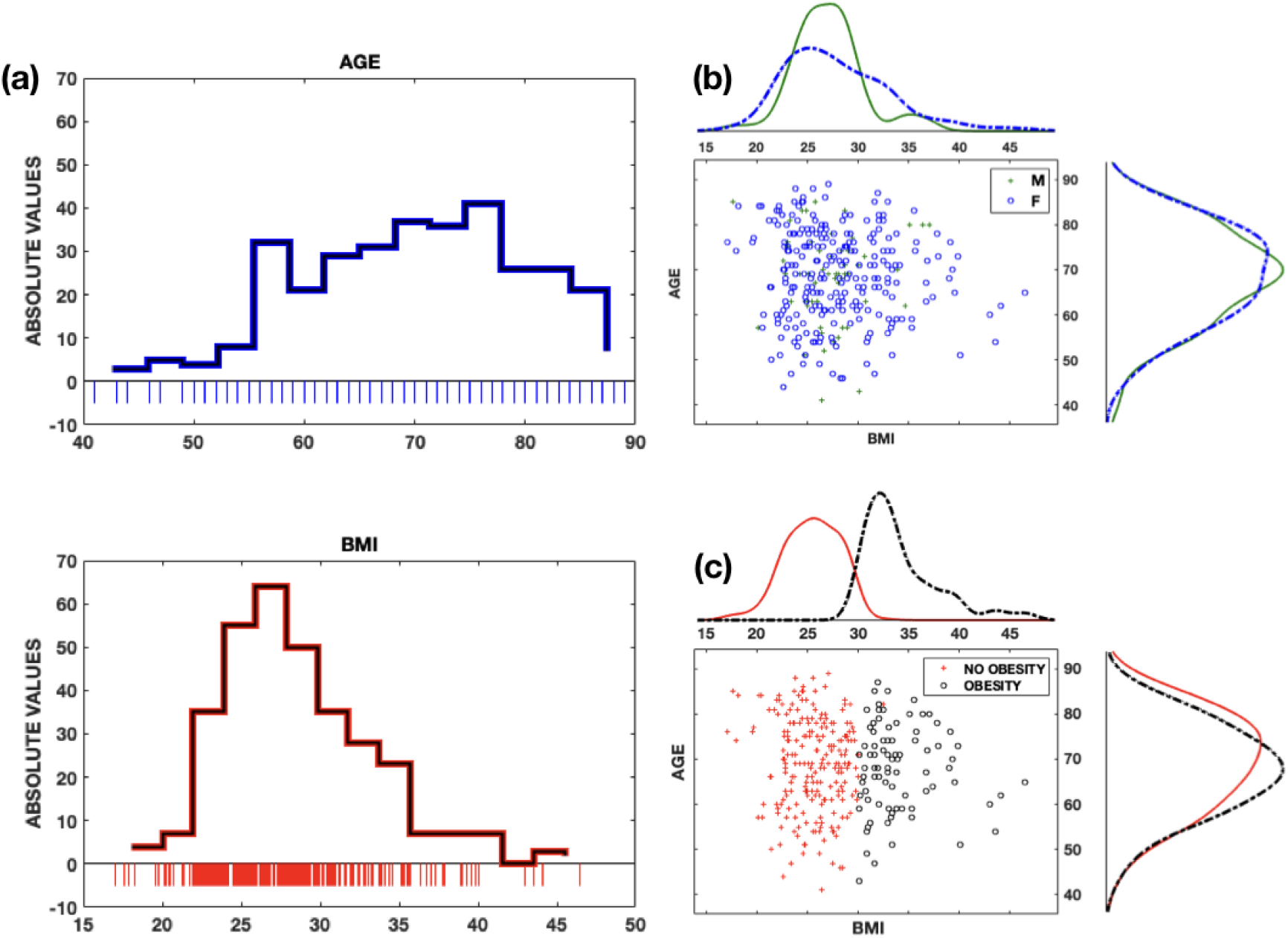
Descriptive analysis of age and BMI distributions. (a) Histogram showing how age (blue) and BMI (red) are distributed on this population of patients with hypothyroidism. The population under study consisted mainly by elderly (68.79 ± 10.05) and no-obese people (27.75 ± 4.84). (b) Scatter plot crossing age and BMI in function of gender showing that there are no differences between these two factors. (c) Scatter plot crossing age and BMI in function of obesity, emphasizing a good balance between these two groups, with a tendency for no-obese people.

The incidence of different comorbidities in this population is described in figure 2 (a). The prevalence of comorbidities is calculated for each age group (major than 60 and minor than 60) separated by gender. It is noted that tabagism is significant different in comparison to the other conditions, for all age classes, except in the class of men older than 60 years. Mainly, for the classes under 60 years, for men and women, the tabagism showed to be differentiated in relation to the other conditions. It is also noted that the prevalence of elderly women (>60) with some comorbidity is higher compared to men. Complementary, figure 2 (b) describes the prevalence of comorbidities enter groups of age given a right comorbidity. The variable INDEP considers the four age/gender groups independent of having a specific comorbidity, but any one of them.

**Figure 2.**
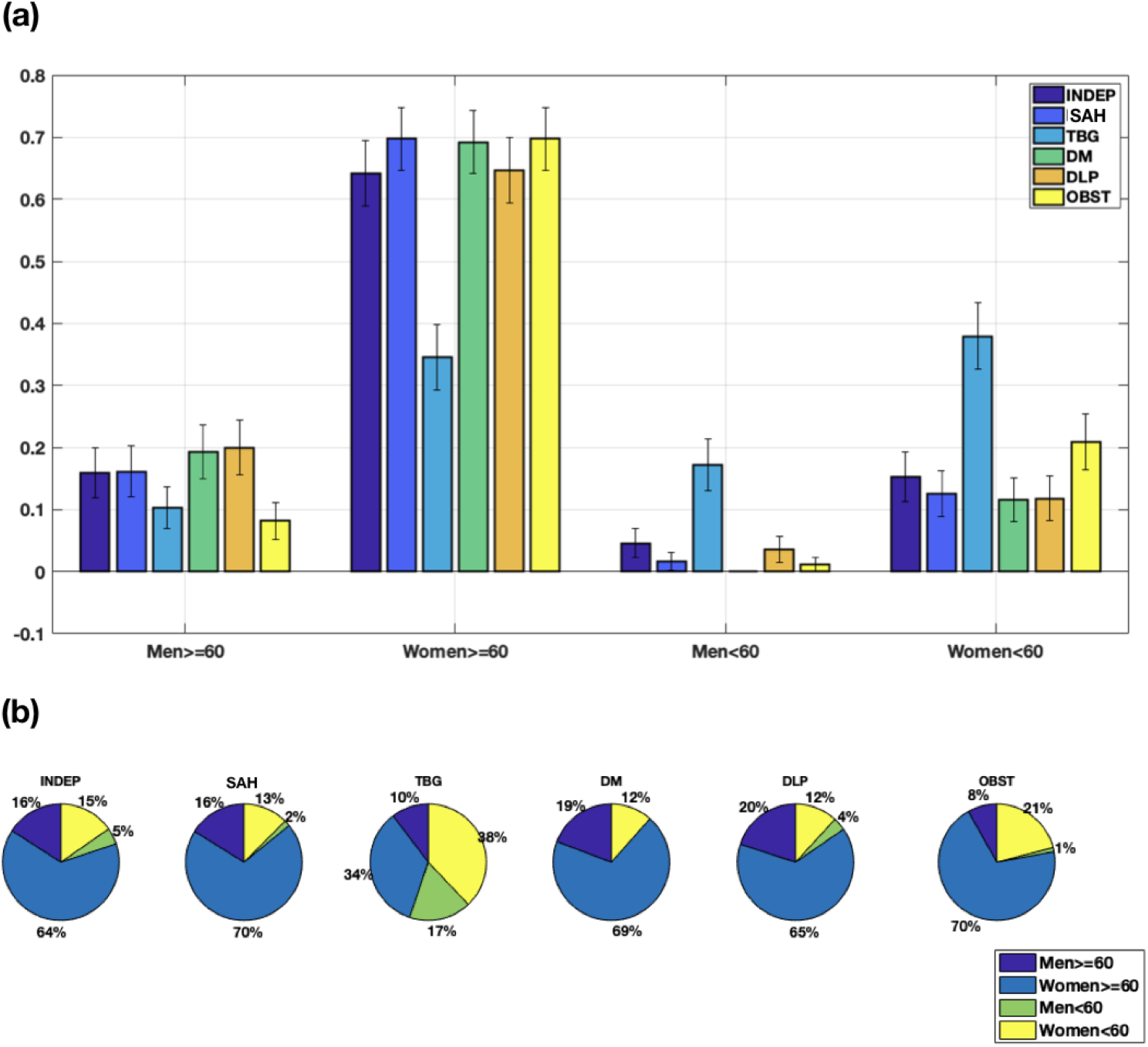
Probabilities associated with each health condition grouped by age. (a) Probabilities of a specific group of age be found with a right condition. Each color highlights the prevalence relative to 5 conditions: systemic arterial hypertension (SAH), tabagism (TBG), diabetes milletus (DM), dyslipidemia (DLP) and obesity (OBST). The term INDEP considers the presence of any one of this 5 conditions, without any distinction. Considering 4 aging groups it is shown that women older than 60 are predominant in this population for all 5 conditions. Tabagism is significant different for all groups but with an inverse tendency for women and men. (b) The same quantification of probabilities is shown, but now with emphasis on the proportion of ages according to each health condition.

To describe the relationship among comorbidities and all possible scenarios for a patient to be diagnosed with MetS, the figure 3(a) shows the distribution of joint probabilities relative to this population. It was considered four scenarios: (i) with no comorbidity (only hypothyroidism), (ii) 1 comorbidity, (iii) 2 comorbidities and (iv) 3 or more comorbidities, separately by age groups. We can observe that there is a higher prevalence of patients with only one comorbidity, mainly for the third and fourth classes (from 60 to 80 years old). The incidence of patients with MetS can be analyzed through the probabilities with three or more comorbidities which are considerably higher than the group with no comorbidity (only hypothyroidism). Additionally, for MetS it includes also the age class from 50 to 60 years old. Since patients with two comorbidities present higher risk to acquire MetS, especially the elderly population (>60), we calculate the probability of these patients to get another comorbidity and increase their chance to be diagnosed with MetS (considering the set of four evaluated comorbidities in this study). Furthermore, for each case of simultaneous comorbidities (i-iv) the age class from 60 to 80 was prevalent in comparison to all others, including to 70 to 80.

**Figure 3.**
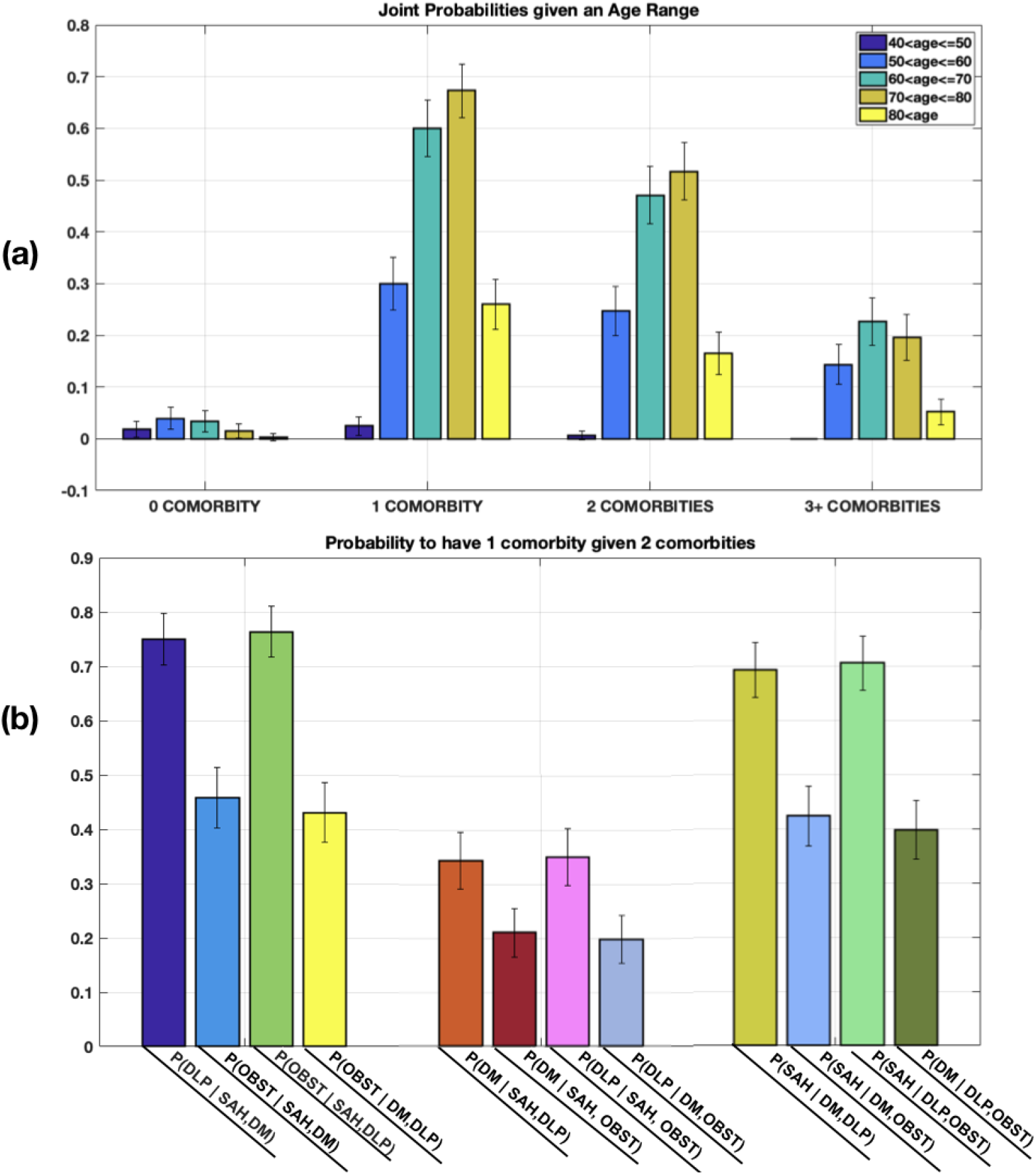
Joint and conditional probabilities for MetS groups of risk. (a) Joint probabilities showing the chances of finding a patient with 0, 1, 2 and 3+ comorbities, distinguished by age classes. Once again patients older than 60 years have more chances to be diagnosed with one or more comorbity, been the main group of risk for MetS. (b) Conditional probability associated with the group that was classified with two comorbities. The plots show the probability of a patient to be diagnosed with MetS since it calculates the probability of acquiring a new disease given that he/she is already with two others.

In Figure 3(b) the conditional probabilities are based on the possibility of acquiring a third comorbidity, considering that the patient already has two comorbidities (higher risk condition).

Figure 3(b) shows that if a patient already has the combination of SAH and DM, he/she has a probability of 72% to get DLP and a probability of 48% to get OBST. This combination, therefore, is one of the highest risks to get MetS. Additionally, when a patient already has the combination of SAH and DLP, he/she has a probability of 76% to get OBST and 34% to get DM. When they have DM and DLP combined, they have a probability of 43% to get OBST and 69% to get SAH. And If patients have SAH and OBST, the chance to acquire DM or DLP are, respectively, 21% and 35%, indicating that these conditions are less likely to occur and develop MetS. Patients that already have DM and OBST comorbidities will have a probability of 20% to get DLP and 42% to get SAH. Finally, if patients already have the combination of DLP and OBST they will have a probability of 70% to get SAH and 40% to get DM. The combination of SAH and DM is, therefore, the worst combination to get MetS.

## 4. Discussion

The statistical survey used a sample comprising of 327 patients with hypothyroidism in which 80% were aged above 60 years and 68% were overweight; in addition, the sample included 75.84% patients with hypertension, 59.63% with dyslipidemia, and 24.16% with diabetes mellitus from a urban region of Brazil (Itajubá city and vicinity). The presence of different comorbidities related to the data, allowed the study of different scenarios associated with MetS and with more risks to get MetS, considering the common occurrence of hypothyroidism for all subjects.

In this sample, we verify that 64% of the patients were women aged above 60 years, figure 2 (b). This data is in accordance with the American Academy of Family Physicians that recommends patients with more than 60 years, mainly women, screening routines in order to track the possibility of hypothyroidism (17).

Figure 1 indicates that, independent of the age, BMI was a relevant factor for this sample. Furthermore, it shows that patients with hypothyroidism are more prone to gain weight, where approximately 75% of the patients have a BMI higher than 25, with a higher concentration of patients with IMB between 25 and 35, featuring a state of overweight patients. Most probably, these conditions are prevalent due to changes in thyroid hormones, which in turn, may worsen or facilitate the presence of other conditions in patients with hypothyroidism (21).

Systemic arterial hypertension, dyslipidemia, diabetes mellitus (DM) and obesity show to be probable risk factors for metabolic syndrome (MetS), in patients with hypothyroidism in this sample. Therefore, since thyroid hormones are crucial in the regulation of glucose and lipid metabolism, they may also interfere with the MetS parameters, such as HDL-cholesterol, thyroglobulin, blood pressure, and plasma glucose (21, 22). That is, Hypothyroidism had a high influence on the chances of having a set of comorbidities related to MetS in this sample, which can contribute to the development of this syndrome in the long term (23).

The comorbidities related to insulin resistance and BMI above the normal range are also considered to be subjacent to MetS since they increase the chances of its diagnosis (23,24). As presented in Figure 3 (a), the percentage of patients that can be already considered with MetS are: 14.37% (between 50 until 59 years), 22.63% (between 60 until 69 years), 19.57% (between 70 until 79 years) and 5.2%(over 80 years) of the studied population. But we also highlight the group of patients with two comorbidities, it represents the group with highest risk of MetS in studied sample.

By comparing all different pairs associated with the conditional probabilities, considering each comorbidity studied in evaluated sample, we calculate the chances of a patient to be diagnosed with a new comorbidity given that he/she already has two others. It was observed that individuals with hypertension (SAH) and diabetes (DM), have a higher chance to acquire dyslipidemia (DLP) than obesity. Other studies also show that there is a greater chance of hypertensive and diabetic patients acquiring DLP, since it is a well-established risk factor for the development of cardiovascular diseases (24, 25). The chance of a patient to be overweight may be associated with his/her tendency to acquire new comorbidities, excepting hormonal diseases, such as hypothyroidism (26). Our data also show that the chance of obese patients (OBST) and those with dyslipidemia (DLP) to acquire diabetes mellitus (DM) is higher than obese patients with hypertension. Most probably it occurs due to the risk factors associated with the simultaneity of comorbidities (26).

In addition, the age factor indicates a trend in elderly patients, mainly systemic arterial hypertension and diabetes mellitus, which increases the chances of developing new comorbidities due to the natural cycle of the disease. Other factors, which are generally associated with hypothyroidism, such as oxidative stress, chronic inflammation, and angiogenesis, demonstrate that they increase the pathogenesis of the metabolic syndrome, but have not been explored in this study (19). Other studies have indicated that some subtypes of hypothyroidism may be associated with the development of MetS and demographic characteristics may alter the comorbid relationships, depending on the criteria adopted as reference values for diagnosis (27).

## 5. Conclusion

Metabolic syndrome is a complex disorder including several cardiovascular risk factors. In this study, we identify, in a sample of patients from a Brazilian urban region, that the chances to develop MetS was considerably high when considering the existence of hormonal basal deficiency, caused by the hypothyroidism. We conclude that age can be a critical factor to develop MetS, mainly between 60 to 80 years. Additionally, the association of hypertension and diabetes (SAH and DM) showed to be the worst combination to influence the development of MetS, followed by DLP and OBST.

## Data Availability

All data was collected by the clinicians and researchers from Medical Faculty of Medicine of Itajuba, at the University Hospital (ASSOCIACAO DE INTEGRACAO SOCIAL DE ITAJUBA).
All the data are accessible under request.

## References

(1) L. K. Meher, S. K. Raveendranathan, S. K. Kota, J. Sarangi, J., S. N. Jali (2013). Prevalence of hypothyroidism in patients with metabolic syndrome. Thyroid Research and Practice, 10(2), 60.

(2) M. T. Nunes, “Hormônios Tiroideanos: Mecanismo de Ação e Importância Biológica,” Arq. Bras. Endocrinol. Metabol., vol. 47, pp. 639–643, 2003.

(3) M. Uzunlulu, E. Yorulmaz., A. Oguz, A. (2007). Prevalence of subclinical hypothyroidism in patients with metabolic syndrome. Endocrine journal, 54(1), 71–76.

(4) A. Gonçalves, E. S. Resende, M. Luiza, M. P. Fernandes, and A. Martins, “Influência dos Hormônios Tireoidianos sobre o Sistema Cardiovascular, Sistema Muscular e a Tolerância ao Esforço: uma Breve Revisão,” Arq. Bras. Cardiol., vol. 87, pp. 5–7, 2006.

(5) J. H. Romaldini and C. S. Farah, “Disfunções Mínimas da Tiróide: Hipotiroidismo Subclínico e Hipertiroidismo Subclínico,” Arq. Bras. Endocrinol. Metabol., vol. 48, no. 1, pp. 147–158, 2004.

(6) G. B. Barra and F. A. R. Neves, “Mecanismo Molecular da Ação do Hormônio Tireoideano,” Arq. Bras. Endocrinol. Metabol., vol. 48, pp. 25–39, 2004.

(7) G. A. De Carvalho, C. Luhm, S. Perez, and L. S. Ward, “The clinical use of thyroid function tests,” Arq. Bras. Endocrinol. Metabol., 2013.

(8) M. Coceani, G. Iervasi, A. Pingitore, C. Carpeggiani, and A. L. Abbate, “Thyroid Hormone and Coronary Artery Disease?: From Clinical Correlations to Prognostic Implications,” Clin. Cardioology, vol. 385, pp. 380–385, 2009.

(9) O. M. Jr, “Hypothyroidism in coronary heart disease and its relation to selected risk factors,” Vasc. Health Risk Manag., vol. 2, no. 4, pp. 499–506, 2006.

(10) A. P. Brandão, A. A. Brandão, A. da R. Nogueira, H. Suplicy, J. I. Guimarães, and J. E. P. de Oliveira, “I Diretriz Brasileira de Diagnóstico e Tratamento da Síndrome Metabólica,” Arquivos Brasileiros de Cardiologia. 2005.

(11) F. Bartoli, G. Carra, and C. Crocamo, “Metabolic Syndrome in People Suffering from Posttraumatic Stress Disorder?: A Systematic Review and Meta-Analysis,” Metab. Syndr. Relat. Disord., vol. 11, no. 5, pp. 301–308, 2013.

(12) G. Brenta, M. Vaisman, J. A. Sgarbi, L. M. Bergoglio, Andrada, N. C. D., P. P. Bravo, H. Graf, H. (2013). Diretrizes clínicas práticas para o manejo do hipotiroidismo. Arquivos Brasileiros de Endocrinologia & Metabologia, 57(4), 265–291.

(13) S. M. Grundy, H. B. Brewer, J. I. Cleeman, S. C. Smith, C. Lenfant, and C. Participants, “Definition of Metabolic Syndrome,” Circ. Am. Hear. Assoc., pp. 433–438, 2004.

(14) S. M. Grundy, D. Becker, L. T. Clark, R. S. Cooper, and M. A. Denke, Detection, Evaluation, and Treatment of High Blood Cholesterol in Adults (Adult Treatment Panel III)’, no. 02.

(15) Erdogan, M., Canataroglu, A., Ganidagli, S., & Kulaksizoglu, M. (2011). Metabolic syndrome prevalence in subclinic and overt hypothyroid patients and the relation among metabolic syndrome parameters. Journal of endocrinological investigation, 34(7), 488–492.

(16) Ruhla, S., Weickert, M. O., Arafat, A. M., Osterhoff, M., Isken, F., Spranger, J., … & Möhlig, M. (2010). A high normal TSH is associated with the metabolic syndrome. Clinical endocrinology, 72(5), 696–701.

(17) K. Chugh, S. Goyal, V. Shankar & Chugh, S.N. (2012). Thyroid function tests in metabolic syndrome. Indian journal of endocrinology and metabolism, 16(6), 958.

(18) B. Rosner, B. (2015). Fundamentals of biostatistics. Nelson Education.

(19) S. K. Kota, L. K. Meher, S. Krishna, K. Modi. (2012). Hypothyroidism in metabolic syndrome. Indian journal of endocrinology and metabolism, 16 (Suppl 2), S332–3.

(20) Rosner, B. (1995). Fundamental of Biostatistics Duxbury Press, Boston.

(21) J. R. Garber, R. H. Cobin, J. R. Garber, and R. H. Cobin, “ATA / AACE Guidelines Clinical Practice Guidelines for Hypothyroidism in adults: cosponsored by the American Association of Clinical Endocrinologist and the American Thyroid Association,” Endocr. Pract., vol. 18., no. 6., pp. 988– 1028, 2012.

(22) M. Cerbone, D. Capalbo, M. Wasniewska et al., “Cardiovascular risk factors in children with long-standing untreated idiopathic subclinical hypothyroidism,” J. Clin. Endocrinol. Metab., vol. 99, no. 8, pp. 2697–2703, 2014.

(23) S. Mahjoub and J. Masrour-Roudsari, “Role of oxidative stress in pathogenesis of metabolic syndrome,” Caspian Journal of Internal Medicine, vol. 3, no. 1, pp. 386–396, 2012.

(24) L. de Pinho, A. P. S. Aguiar, M. R. Oliveira, N. A. P. Barreto, C. M. M. & Ferreira (2015). Hipertensão e dislipidemia em pacientes diabetes mellitus tipo 2: uma revisão integrativa. Renome, 4(1), 87–101.

(25) C. A. T. Radovanovic, L. A. D. Santos, M. D. D. B. Carvalho, S. S. & Marcon (2014). Hipertensão arterial e outros fatores de risco associados às doenças cardiovasculares em adultos. Revista Latino-Americana de Enfermagem, 22(4), 547–553.

(26) I. N, Queiroz (2014). Obesidade em hipertensos e/ou diabéticos cadastrados na Estratégia Saúde da Família (ESF) Vila São Francisco de Assis, no município de Montes Claros-MG.

(27) L. Yang, X. Lv, F. Yue, D. Wei, W. Liu, and T. Zhang, “Subclinical hypothyroidism and the risk of metabolic syndrome: a meta -analysis of observational studies,” Endocr. Res., vol. 41, no. 2, pp. 158–65, 2016. DOI: 10.3109/07435800.2015.1108332

